# Patterns of Autism Symptoms: Hidden Structure in the ADOS and ADI-R instruments

**DOI:** 10.1101/19007419

**Authors:** Jérémy Lefort-Besnard, Kai Vogeley, Leonhard Schilbach, Gaël Varoquaux, Bertrand Thirion, Guillaume Dumas, Danilo Bzdok

## Abstract

We simultaneously revisited the ADI-R and ADOS with a comprehensive data-analytics strategy. Here, the combination of pattern analysis algorithms and an extensive data resources (n=266 patients aged 7 to 49 years) allowed identifying coherent clinical constellations in and across ADI-R and ADOS assessments widespread in clinical practice. The collective results of the clustering and sparse regression approaches suggest that identifying autism subtypes and severity for a given individual may be most manifested in the social and communication domains of the ADI-R. Additionally, our quantitative investigation revealed that disease-specific patterns of ADI-R and ADOS scores can be uncovered when studying sex, age or level of FIQ in patients.

## INTRODUCTION

Autism spectrum disorder is clinically defined by different types of symptoms: deficits in communication and social interaction as well as restricted, repetitive patterns of behavior, interests, or activities. It is a spectrum disorder because a patient’s symptoms can present as a wide variety of combinations, from mild to severe. It is yet not clear how specific subgroups may map onto symptom presentations (Frith & Happé, 2005).

The clinical assessment of autism symptoms is still a lengthy and time-consuming process. This evaluation involves a team of well-trained professionals and combines information about the patient from different sources. The assessment of such patient includes an interview with the caregivers as well as an observation of each individual both in the living environment and as part of a psychiatric evaluation in the assessment center. Two clinical tools have become established in this assessment procedure and are considered to be the ‘gold standard’ in symptom evaluation for autism, particularly when combined with clinical judgment: i) the ADI-R (Lord, Rutter, & Le Couteur, 1994), a semi-structured interview conducted with parents which focuses on current presentation and lifelong developmental history, and ii) the ADOS (Lord et al., 2000), a standardized semi-structured diagnosis assessment conducted through one-to-one personal interaction and direct observation of an individual suspected to have autism using a range of activities and delivered by a trained examiner. A combination of ADI-R and ADOS assessments coupled with experienced clinical judgments has previously been shown to improve diagnostic validity (Kim & Lord, 2012). In everyday clinical practice, however, often only one of the instruments is used due to time, cost or expertise constraints. The choice of preferred instrument used for the evaluation of autism symptoms remains quite variable from one center to another.

The inconsistency in the choice of symptom assessment tool used might have contributed to discrepancies in results and conclusions of several studies (Grantham et al., 2014; Lemler, 2012). Only a handful of studies have directly compared the ADI-R and ADOS and the outcomes are not always directly clinically applicable. For instance, De Bildt and colleagues (2004) found that the level of agreement between the ADI-R and ADOS was slightly higher than the chance level to classify individuals as patients with autism, patients with pervasive developmental disorder or healthy. In addition, Mazefsky and Oslwald (2006) found a good agreement between team diagnoses and the instruments. As such, the commonalities and divergences between the widespread ADI-R and the ADOS are yet to be fully understood.

The use of these two symptom assessment tools required long training and each of them is rather time-consuming. These circumstances can lead to massive delays in diagnosis and unequal coverage of the population in need of medical attention (Goin-Kochel, Mackintosh, & Myers, 2006). As a consequence, delays in the delivery of therapies occur frequently, which contributes to parental distress (Goddard, Lehr, & Lapadat, 2000; Quine & Pahl, 1987) and may also affect the long-term outcomes of early interventions (Goin & Myers, 2004). For these reasons, it is an emerging agenda to improve these procedures to diminish the time between symptom appearance and formal diagnosis. Indeed, streamlining the process to detect autism severity in a rapid, efficient, and accurate fashion could lead to significant benefits. Such gains would include saving patients time and potentially allowing for fast early intervention, reduction of clinician work hours and alleviating economic costs to name a few.

In view of potential benefits from refining the diagnostic process and the need of a more direct comparison of each instrument, we simultaneously revisited the ADI-R and ADOS with a comprehensive data-analytics strategy. Variability in symptoms and severity within the autism population suggests the possibility of dimensional profiles of autism symptoms. Therefore, we extracted distinct homogeneous patient symptom profiles from the ADI-R and ADOS giving further insights on how symptoms presentation might outline different patient subgroups. Additionally, we used data-guided evaluation for the parsimonious subsets of the ADI-R and ADOS which could enable improvements of clinical workflows. Finally, we quantitatively characterized the relation between the two instruments in the context of different patients’ age, sex and fluid IQ (FIQ).

## METHODS

### Data resources

We systematically charted the relationship between the ADI-R and the ADOS based on behavioral data from a publicly available dataset: ABIDE (Autism Brain Imaging Data Exchange; http://fcon_1000.projects.nitrc.org/indi/abide/). All data analyses were carried out on this data repository which is described in detail in Di Martino and colleagues (2014). The ABIDE data provides subject information, including age, sex, measures of intellectual functioning, and measures of symptom severity as assessed by the Autism Diagnostic Observation Schedule (ADOS) as well as the Autism Diagnostic Interview-Revised (ADI-R). The behavioral assessments were collected from a total of 266 patients with an autism spectrum disorder diagnosis, aged 7/49 years, including 233 male and 33 female subjects (see Supplementary Table 1 for details). The autism diagnoses were provided by board-certified psychiatrists. The distribution of the responses in ADI-R and the ADOS assessment in our sample was homogeneous (SFig. 1). Homogeneous distribution of the ADI-R and ADOS scores was also found across patients’ age (SFig. 2) The original studies included in ABIDE received approval from each site’s Institutional Review Board.

### Identifying hidden group structure: k-means clustering

To explore distinct subgroups among patients with autism, we applied a k-means clustering algorithm to automatically partition patient symptom profiles into homogeneous groups. K-means is a method identifying many-to-one mapping (Forgy, 1965): each patient is a member of exactly one group. We used “NbClust” (Charrad, Ghazzali, Boiteau, Niknafs, & Charrad, 2014) as an established R package to simultaneously apply 30 cluster validity metrics. This approach provided complementary ways to reach indications of the number of groups most supported by the patient data. Among all metrics of cluster usefulness and according to the majority rule, the best number of clusters was three (see the supplementary table 2 for the output of the NbClust package in R). That is, the most robust number of patient symptom clusters consisted of three groups, to the extend supported by our data. Therefore, three patient groups of distinct symptom profiles were automatically extracted as it provided a useful fit to our clinical sample.

### Identifying predictive relevance of domains of the ADI-R and ADOS: Sparse logistic regression

The goal behind k-means was to partition the patients into non-overlapping homogeneous groups (Bzdok & Meyer-Lindenberg, 2017) as measured by the ADI-R and ADOS domains. This approach allowed to explore the relationship among autism spectrum disorder patients. Complementing these insights in a next step, we applied a modeling technique that emphasizes prediction performance with an optimal tradeoff against the number of most relevant domains.

To extract the most informative subsets of domains for predicting autism severity, we capitalized on the pattern analysis algorithm *sparse logistic regression* (Hastie, Tibshirani, & Wainwright, 2015). The sparsity constraint was imposed in form of an *l*_1_ regularization penalty added to the objective of the generalized linear modelling problem. Such a constraint in the optimization objective automatically detects relevant features “on-the-fly” during model estimation. The*l*_1_ penalty term, calibrated by the hyper-parameter λ, exerts control over the parsimony criterion and its shrinkage regularization on the learned model weights. The penalized negative log-likelihood of the sparse logistic regression objective is given by:

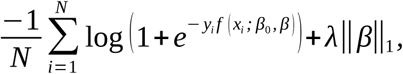

where *x*_*i*_ represents a given patient’s ADI-R and ADOS domain scores, *y*_*i*_ is his or her autism severity group defined as the median-split of the ADI-R and ADOS total score (i.e., 0 as mild, 1 as severe) representing a categorical summary of the constituent continuous scores, *β*_0_ is the intercept, and *β* is the weight attached to each instrument domain, the right term corresponds to the *l*_1_ penalty term controlled by the hyper-parameter λ. The domain selection behavior is calibrated by the choice of this tuning parameter. The hyper-parameter selection was based on an independent split of our data in a principled fashion using nested cross-validation (X data splits in each fold). In a common grid of candidate tuning parameter choices, the value of λ was varied logarithmically from 3.5 to 1.0 in log-space with 16 steps. The member in the model family that yielded highest prediction accuracy (i.e., generalization performance) for each candidate of λ was selected. In other words, the goal here was not to select the best hyper-parameter. Rather, we charted a space of candidate λ to explicitly investigate the parsimony tradeoff from imposing high to low sparsity. In this way, the quantitative investigation detected subsets of domains that were most informative about the autism severity.

### Assessing domain importance in predicting autism severity

The k-means method (cf. above) extracted latent structure dormant in the data regardless of symptom severity measures. Sparse logistic regression (cf. above) in turn selected the most predictive variables. Here, we wanted to make sure that the sparse logistic regression was more appropriate to our research setting than a model looking for higher-order effects. We thus compared the prediction performance of the sparse logistic regression to the accuracy reached by a commonly used non-linear predictive model: the random-forest algorithm.

### Predicting age, sex, and FIQ based on the ADI-R and ADOS domains

We analyzed the relative importance of each ADI-R and ADOS domain for predicting sex, age and FIQ using a logistic regression (without sparsity constraint). Note that we refrained from computing more sophisticated regressions with non-linear interaction terms because these model extensions would require more than twice the sample size to obtain model fits of equal quality. Subgroups representative of each category were extracted. We then looked for the weight of each instrument domain to predict the outcome of interest. The age and FIQ outcomes were defined as categorical summaries of the constituent continuous scores. That is, the patients’ age was defined as superior or inferior to 18 years, while the patients’ level of FIQ was defined as higher or lower than the median-split of the FIQ scores (105.35). Note that the FIQ was used as measure of interest given the high number of missing entries in the other IQ measurements provided in the ABIDE dataset.

In a first set of analyses, the sample was subdivided based on FIQ and sex. That is, four subgroups were extracted depicting males with high FIQ, males with low FIQ, females with high FIQ and females with low FIQ. A logistic regression was applied to predict the age based on the domain scores of the ADI-R and ADOS of each specific subgroup.

For the second set of analyses, the sample was subdivided based on FIQ and age. Thus, we ended up with four subgroups including respectively adults with high FIQ, adults with low FIQ, teenagers with high FIQ and teenagers with low FIQ. A logistic regression was applied to predict the sex based on the scores of the ADI-R and ADOS domains of each specific subgroup.

For the third set of analyses, the patient sample was subdivided based on sex and age. This subdivision led to four subgroups including respectively adult males, adult females, teenager males and teenager females. A logistic regression was applied to predict the FIQ based on the scores of the ADI-R and ADOS domains of each specific subgroup.

For each of these analyses, in each subgroup, the logistic regression was applied only if more than 10 observations were available. Furthermore, class imbalance, if present, was handled using up-sampling if the majority class was inferior or equal to a third of the minority class or using down-sampling otherwise. Once the weights of the logistic regression for a subgroup computed, the 90% bootstrapped confidence intervals were calculated by fitting the logistic regression to 100 bootstrapped samples made from the specific subgroup. In a similar fashion as described above, bootstrapping was used here to provide a principled estimate of the statistical quality of the domain relevances for estimating age, sex and FIQ. Finally, we computed the confusion matrix for each classification algorithm to allow visualization of its performance.

### Code availability

All analysis scripts of the present study are readily accessible to the reader online (https://github.com/JLefortBesnard/ADIR_ADOS2019). See Supplementary Methods for more details.

## RESULTS

### Properties of patient groups hidden in the ADI-R and ADOS assessments

To explore distinct subgroups related to the ADI-R and ADOS assessment patterns among patients with autism, we assigned each patient to one dominant symptom constellation based on the two instruments. This data-driven exploration revealed three distinct symptom constellations that grouped the patients in our sample (Fig. 1): i) A severe group that included patients scoring high on every domain of the ADI-R and ADOS. Specifically, these patients had on average particularly high scores on the domains of the ADOS. ii) A mild group that included patients who scored low in every domain of the ADI-R and ADOS. By contrast, this coherent subgroup of patients with similar profiles exhibited particularly low scores on the social and communication domains of the ADI-R. iii) An ADOS-negative group that included patients who scored high in every domain of the ADI-R and low in every domain of the ADOS. In sum, the divergence between the social and communication domains as assessed by the ADI-R and the ADOS were the most informative markers. In contrast, the least instructive marker was the repetitive behavior domain of the two instruments (SFig. 3). The three subgroups were homogeneous in terms of sex, age and FIQ (STable 3).

**Figure 1.**
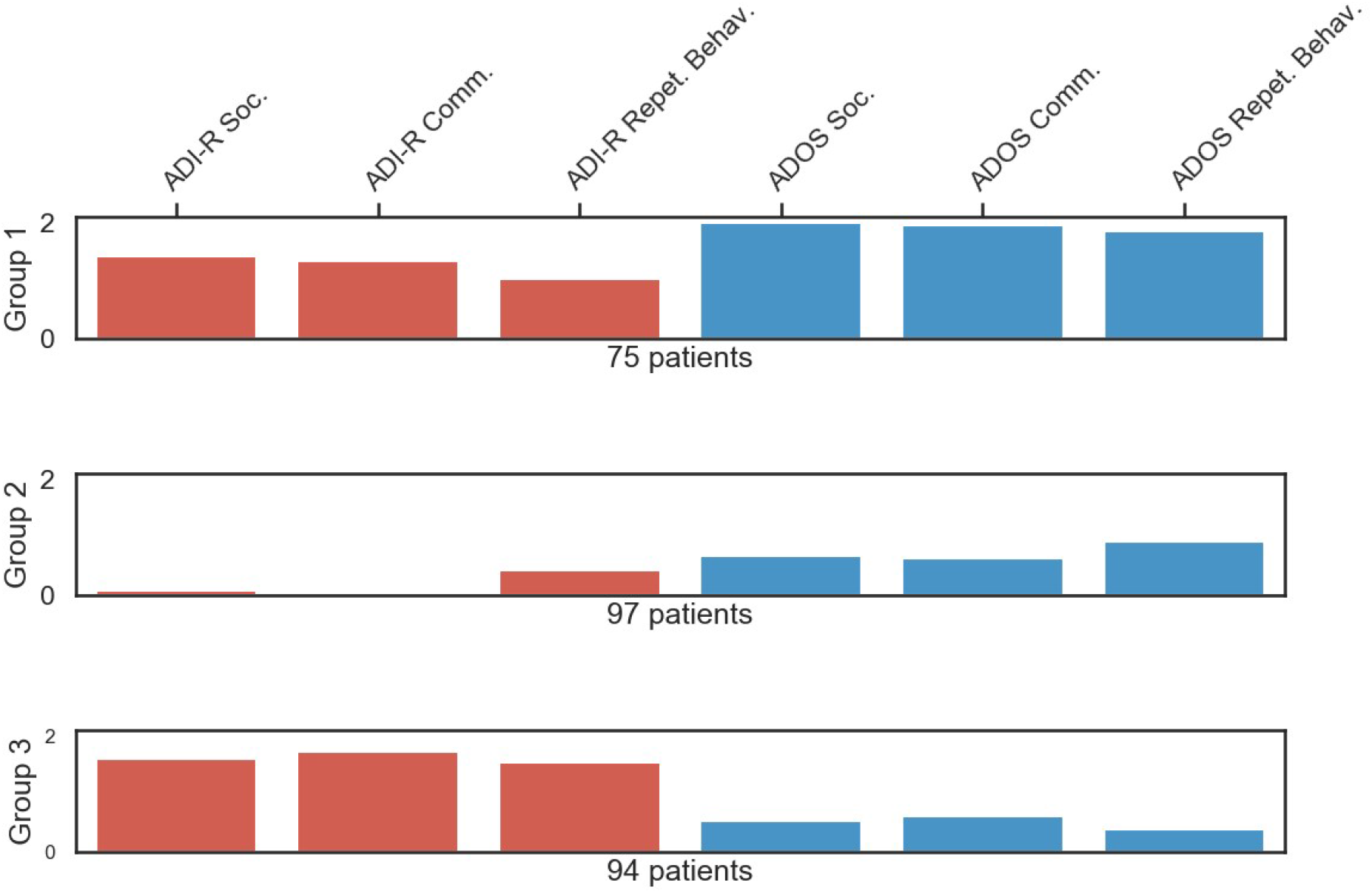
Uncovering three patient groups with distinct symptom profiles. Automatic clustering based on the domains of the ADI-R and ADOS questionnaires exposed three distinct symptom constellations that grouped the patients. Each row represents one data-derived symptom group constellation hidden in the patient assessments. The weights (*y axis*) of each domain (*x axis*) indicated the relative importance of the domains for a particular group in the k-means model. The *red* bars are the domain scores of the ADI-R in the respective cluster and the *blue* ones are the domain scores of the ADOS. Three different autism subtypes emerged: i) a *severe* profile including patients scoring high on every domain of the ADI-R and ADOS with particularly high scores on the domains of the ADOS (*group 1*), ii) a *mild* profile including patients scoring low in every domain of the ADI-R and ADOS with particularly low scores on the social and communication domains of the ADI-R (g*roup 2*), and iii) an *ADOS-negative* profile including patients scoring high in every domain of the ADI-R and scoring low in every domain of the ADOS (g*roup 3*). These findings from automatic clustering provide additional evidence that the ADI-R and ADOS questionnaires capture some distinct clinical aspects of patients with autism. Furthermore, our results suggest that the symptoms severity of the ADOS-negative group might be more accurately measured by the ADI-R.

As an exploratory pattern-discovery approach, k-means yields clusters as descriptive summary of our data, without primary concern for predictive validity (Bzdok, 2017; Shalev-Shwartz & Ben-David, 2014). A natural next step of the present study therefore consisted in estimating the predictability of autism from ADI-R and ADOS instruments domains.

### Isolating the most predictive domains in the ADI-R and ADOS instruments

A sparse logistic regression was used to automatically identify domain subsets in the ADI-R and ADOS instruments that are most informative about telling mild versus severe autism apart in future patients. With systematically varying parsimony constraint, a series of algorithms estimations was carried out to predict autism severity (defined as the median-split of the ADI-R and ADOS total score) based on the symptom scales (Fig. 2 A and 2 C).

**Figure 2.**
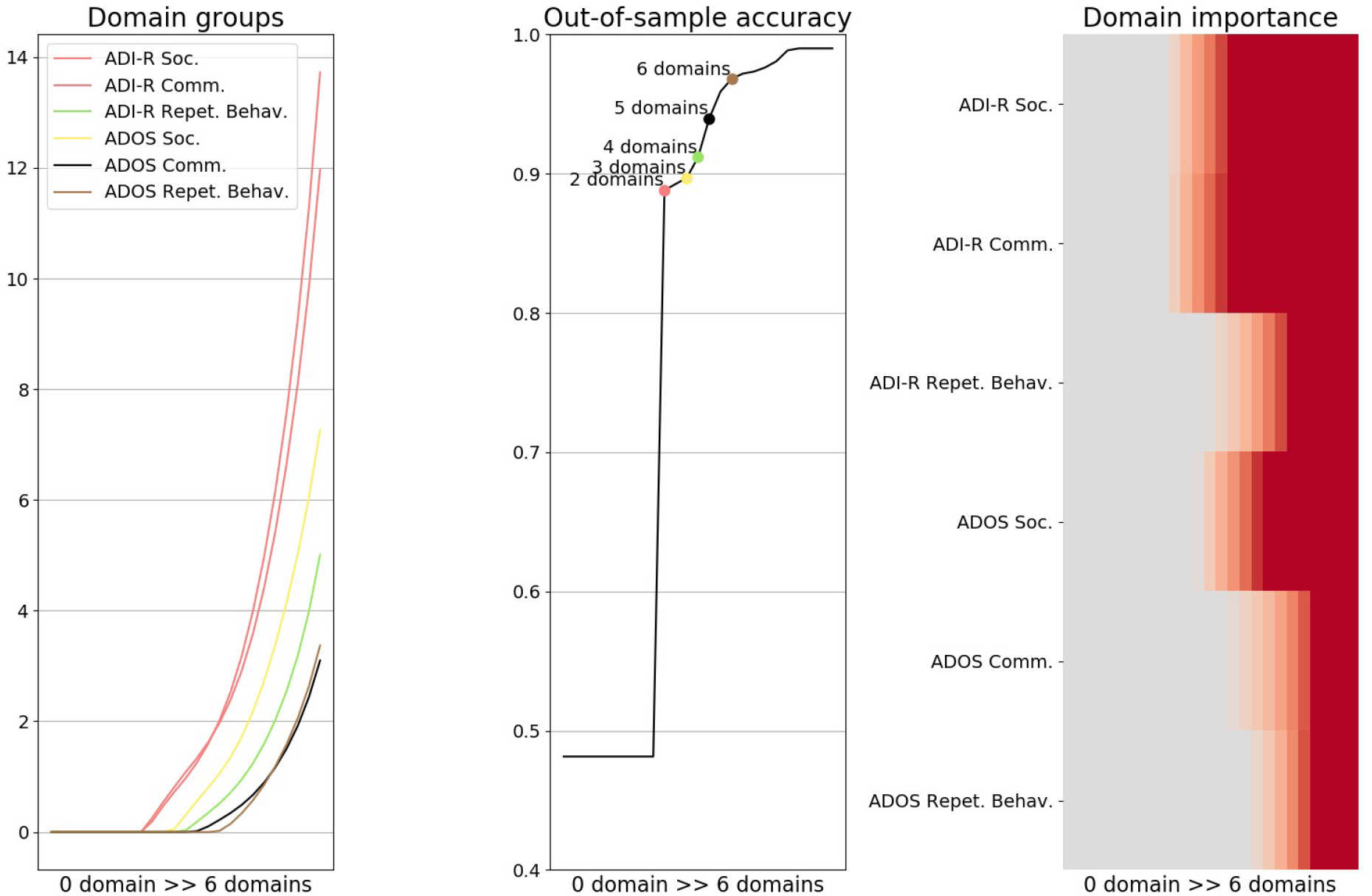
Predictive decomposition of autism symptoms. A parsimony-inducing pattern analysis algorithm was used to search through the array of questionnaire domains and extract the most informative subsets of domains for predicting symptom severity in patients with autism. *A)Domain groups* Trajectories of the classifier weights of the ADI-R and ADOS domains are plotted on the y axis while the parsimony constraint of the statistical models decreasing from left to right (here represented as the increasing number of domains automatically selected) is plotted on the x axis. The curves indicate changes in the subset of selected domains (i.e., the weight set not equal to zero), typically an inclusion. The color of each line shows in which model solution a specific questionnaire domain is included as relevant. For example, ADI-R social and communication domains were found as part of the first (most parsimonious) predictive model and are plotted in red. *B)Prediction accuracy* The middle panel retraces how prediction performance increases step-by-step as the identified domain subsets are added to the model. Each colored point represents a predictive model including a specific number of selected domains. Two domains were sufficient for decent prediction performance at the single-subject level. These two domains predicted autism severity with 88.81% accuracy, while the model including every domains of the ADI-R and ADOS predicted autism severity with 96.81% accuracy. *C)Relative domain importance* Domain importance in the active weights is indicated as the parsimony constraint becomes more lenient (*left to right*). This panel thus represents the relative importance of each domain (*y axis*) as more variables are included in the model (*x axis, from left to right*). In sum, the results emphasize that using every domains of the ADI-R and ADOS, autism severity was predicted with 97% accuracy, while using only the social and communication domains of the ADI-R, autism severity was predicted with an accuracy as high as 89% indicating a very high predictive power for these two elements.

Our analysis strategy extracted two of the overall six domains as the most predictive subset and achieved quite effective prediction of autism severity (88.81% accuracy). This essential subset included the social and communication domains of the ADI-R.

As we calibrated the parsimony constraint step-by-step, four other solutions were automatically identified that isolated further subsets of instrument domains predictive of autism severity (Fig. 2 B). The second solution included the social domain of the ADOS in addition to the social and communication domains of the ADI-R and reached a prediction accuracy of 89.7%. The three remaining automatically identified solutions successively incorporated to the previous subset the repetitive behavior domain of the ADI-R, the communication domain of the ADOS and finally included the repetitive behavior of the ADOS. These three final domain inclusions allowed for a prediction accuracy of 91.18%, 93.96%, and 96.81% in new patients, respectively. In other words, rich descriptions of the patterns in the ADI-R and the ADOS were extracted and two domains of the ADI-R were identified as being highly predictive of autism severity. In sum, though the combination of both ADI-R and ADOS instruments are required for a more rigorous diagnostic procedure, our data-guided analysis gave support to the existence of a subset including two ADI-R domains highly predictive of autism severity.

### Testing for non-linearity in the ADI-R and ADOS instruments

To complement the sparse logistic regression insights, we combined exploration of more sophisticated domain-domain relationships with the evaluation of prediction performance using random-forest algorithm. Random-forest reached a maximum prediction accuracy of 92.78%, while the accuracy obtained with the sparse logistic regression using every domain of the ADI-R and ADOS reached 96.81% (SFig. 4). Interestingly, both the random-forest algorithm and the sparse logistic regression found the social and communication domains of the ADI-R to exhibit patterns of interest in the estimation of autism severity.

### Exploring the instrument domains relation to patients’ age, sex and FIQ

In a first set of analyses, to quantify the relation of the ADI-R and the ADOS domains with their relation to age, we explored the contribution of each instrument domain in deriving age of patients in our sample. *A*ge (i.e., target variable) was predicted based on the ADI-R and ADOS domains (i.e., input variables) after segregating the patient pool into subgroups according to sex and FIQ (Fig. 3, SFig. 5). For this analysis and the following ones, only the domains with important weight per estimation and exhibiting mostly one-sided confidence interval were interpreted (STable 4). In this first analysis, the communication domain of the ADOS contributed to detecting an adult, while the ADI-R repetitive behavior domain contributed to detecting a teenager in each subgroup. In other words, a patient scoring high in the ADOS communication domain would tip the balance of the output toward being an adult, while scoring high in the ADI-R repetition behavior domain would tip it toward being a teenager. In females with high FIQ, the ADI-R communication domain was highly weighted contributing to being a teenager. In males with high FIQ, the social domain of the ADI-R contributed to detecting an adult, while in males with low FIQ, the social domain of the ADOS contributed to detecting a teenager. Finally, the repetitive behavior domain of the ADOS was contributing to being an adult in males and females with high FIQ, while this ADOS domain was contributing to being a teenager in males with low FIQ.

**Figure 3.**
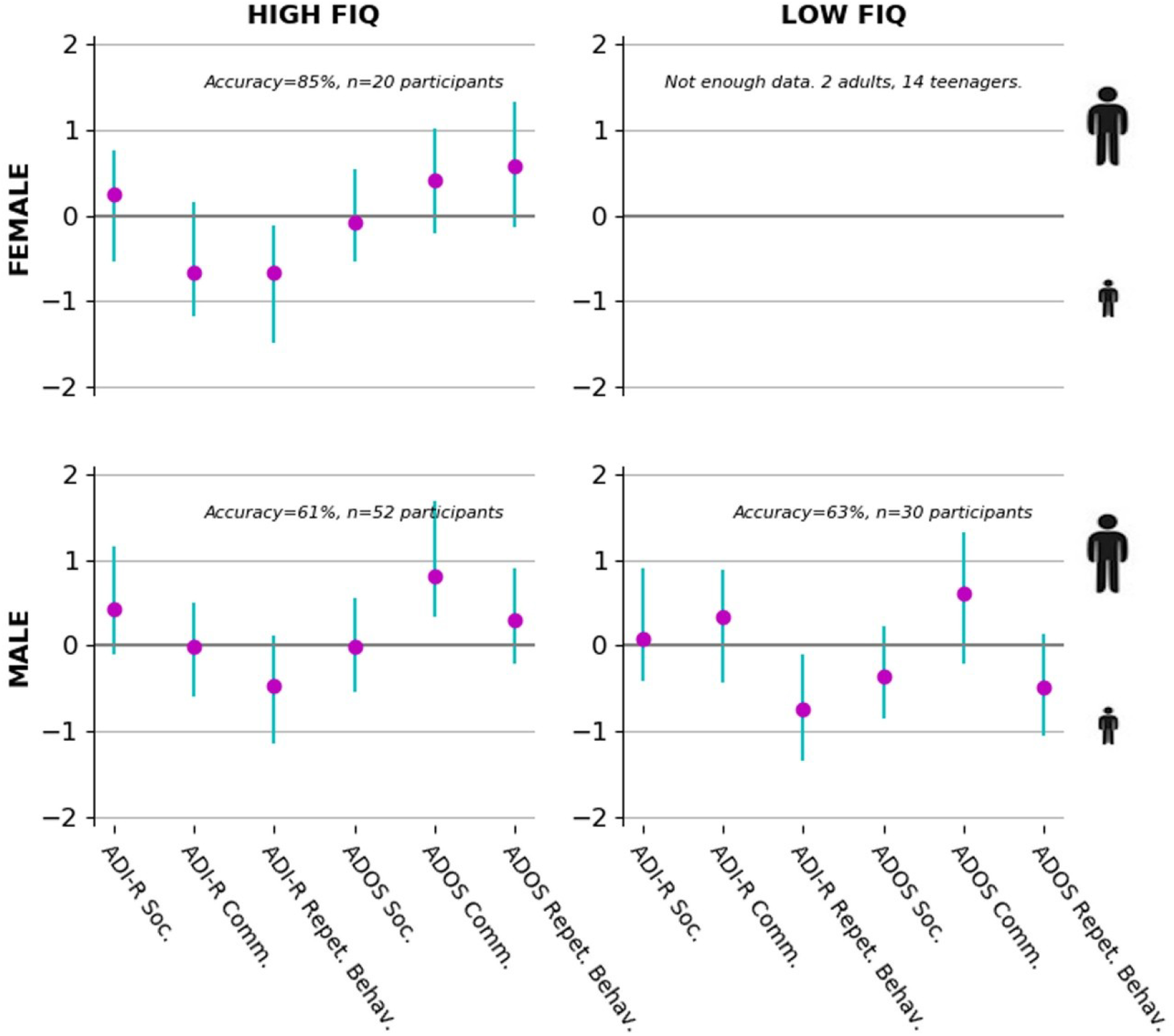
Domain importance in age prediction segregated by sex and FIQ. Adult versus teenager was distinguished based on domains quantified in the ADI-R and ADOS. Each plot shows identical analyses on a stratified subset of the patient pool. From upper left to lower right, we selected the male patients with high or low FIQ, and the female patients with high FIQ. We had insufficient female patients with low FIQ to perform the analysis. The purple circles show the estimated contribution (*y-axis)* of each particular questionnaire domain (*x-axis*) to distinguishing adult versus teenager participants using logistic regression in each specific subgroup. Each green bar indicates the bootstrapped 90% uncertainty interval at the population level. The communication domain of the ADOS contributed to detecting an adult in our sample, while a patient scoring high in the repetitive behavior of the ADI-R was more likely to be a teenager. In other words, a patient scoring high in the communication domain of the ADOS would tip the balance of the output toward being an adult, while scoring high in the repetitive behavior domain of the ADI-R would tip it toward being a teenager. In females with high FIQ, the communication domain of the ADI-R was most associated with being a teenager. In males with high FIQ, the social domain of the ADOS contributed to detecting a teenager, while in males with low FIQ the communication domain of the ADI-R contributed to detecting an adult. Finally, the repetitive behavior domain of the ADOS was contributing to detecting male and female adults with high FIQ, while this ADOS domain was contributing to detecting a teenager in males with low FIQ.

In a second set of analyses, to quantify the relation of the ADI-R and the ADOS domains with their relation to sex, we explored the contribution of each instrument domain in deriving sex of patients in our sample. Sex (i.e., target variable) was predicted based on the ADI-R and ADOS domains (i.e., input variables) after segregating the patient pool into subgroups according to age and FIQ (Fig. 4, SFig. 6). The ADI-R was not contributing for differentiating sex. The ADOS social domain was highly weighted contributing to identifying females among both adult with high FIQ and teenager with low FIQ. The ADOS communication contributed to detecting a male in adult with high FIQ and to being a female in teenagers with high FIQ. The ADOS repetitive behavior domain was highly weighted in teenagers with high FIQ contributing to being a male.

**Figure 4.**
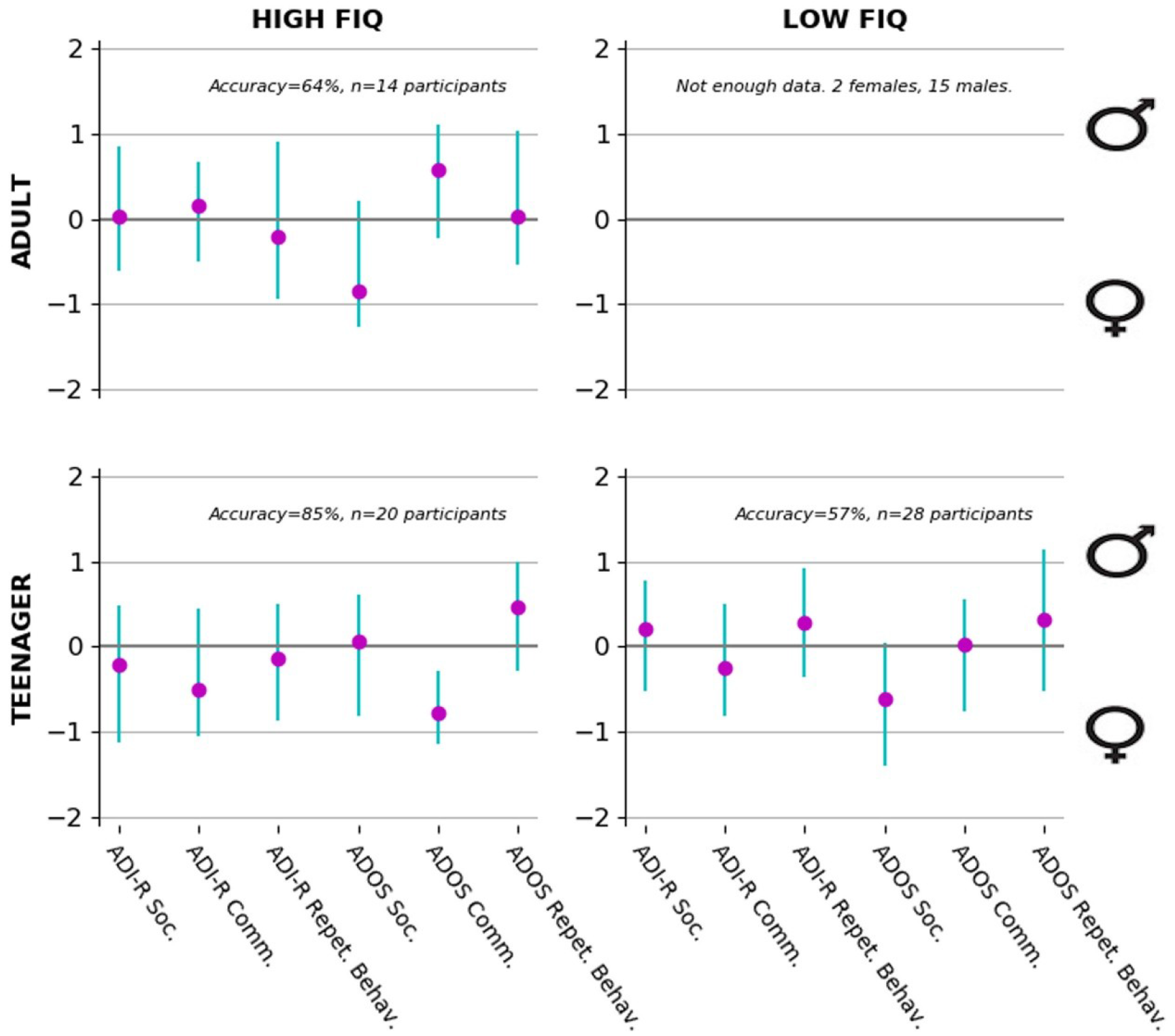
Domain importance in sex prediction segregated by age and FIQ. Male versus female patients were distinguished based on domains quantified in the ADI-R and ADOS. Each plot shows identical analyses on stratified subset of patient pool. From upper left to lower right, we selected the adult patients with high FIQ, and the teenager patients with high or low FIQ. We had insufficient adult patients with low FIQ to compute the analysis. The purple circles show the estimated contribution (*y-axis)* of each particular questionnaire domain (*x-axis*) to distinguishing male versus female participants using logistic regression in each specific subgroup. Each green bar indicates the bootstrapped 90% uncertainty interval at the population level. The ADI-R domains hardly contributed to differentiating sex. The social domain of the ADOS was highly associated with detecting a female in both adult with high FIQ and teenager with low FIQ. The communication domain of the ADOS enabled detecting a male in adults with high FIQ and a female in teenagers with high FIQ. The repetitive behavior domain of the ADOS was highly weighted in teenagers with high FIQ contributing to detecting males.

In a final set of analyses, to quantify the relation of the ADI-R and the ADOS domains with their relation to FIQ, we explored the contribution of each instrument domain in deriving the level of FIQ of patients in our sample. FIQ (i.e., target variable) was predicted based on the ADI-R and ADOS domains (i.e., input variables) after segregating the patient pool into subgroups according to age and sex (Fig. 5, SFig. 7). In teenager males, the repetitive behavior domain of the ADI-R as well as the social and repetitive behavior domains of the ADOS were contributing to having a low FIQ, while the ADOS communication domain contributed to detecting a high FIQ. In adult females, the communication domain of the ADI-R as well as the social domain of the ADOS contributed to detecting a low FIQ, while the repetitive behavior domain of the ADI-R contributed to detecting a high FIQ. In teenager females, the communication and repetitive behavior domains of the ADI-R as well as the communication domain of the ADOS were highly weighted contributing to having a high FIQ.

**Figure 5.**
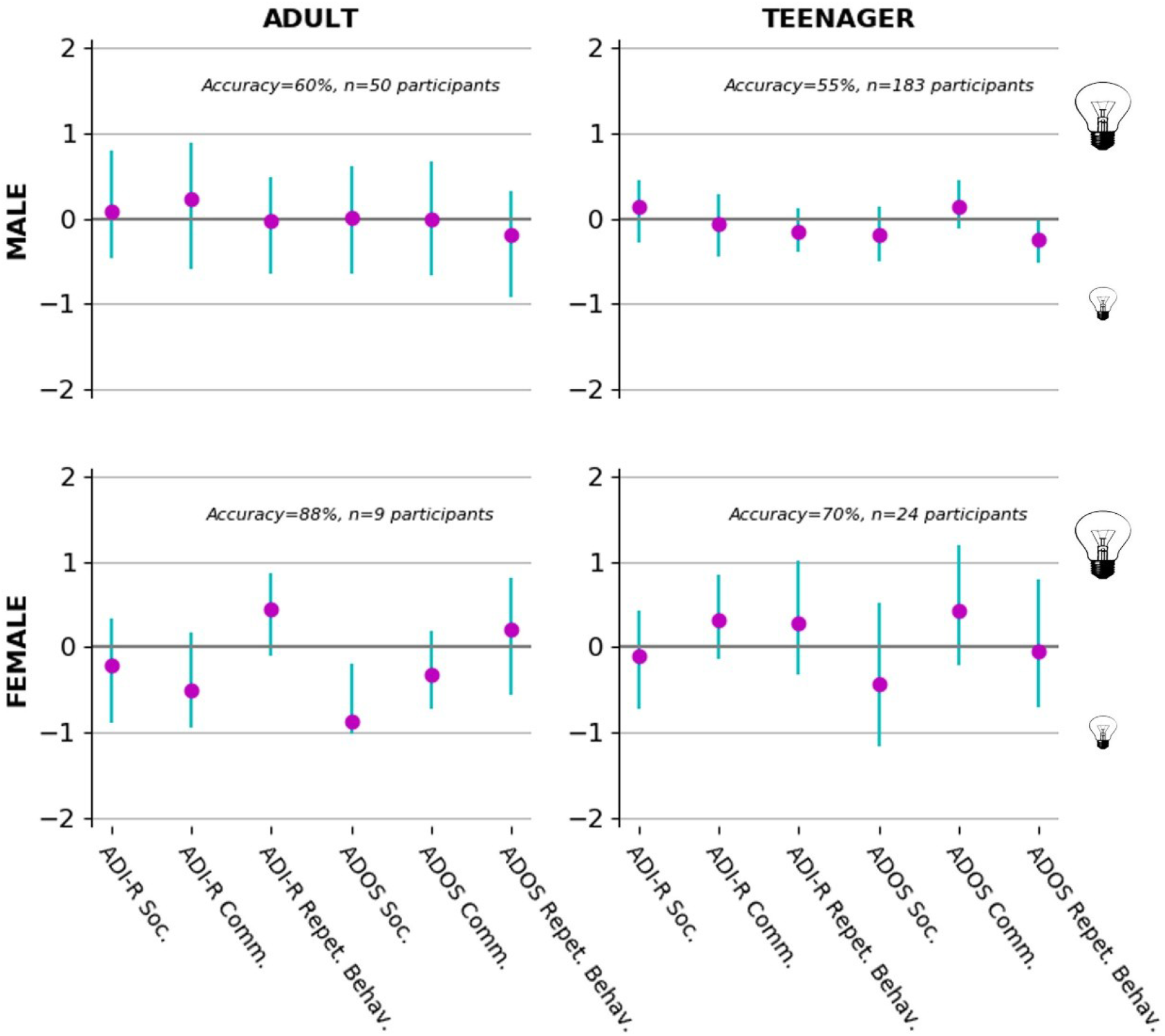
Domain importance in FIQ prediction segregated by sex and age. High versus low FIQ was distinguished based on domains quantified in the ADI-R and ADOS. Each plot shows identical analyses on a stratified subset of the patient pool. From upper left to lower right, we selected the adult or teenager male patients and the adult or teenager female patients. The purple circles show the estimated contribution (*y-axis)* of each particular questionnaire domain (*x-axis*) to distinguishing participants with high versus low FIQ using logistic regression in each specific subgroup. Each green bar indicates the bootstrapped 90% confidence interval at the population level. In teenager males, the repetitive behavior of the ADI-R, the social and repetitive behavior domains of the ADOS were contributing to having a low FIQ, while the communication domain of the ADOS helped detecting high FIQ. In adult females, the communication domain of the ADI-R and the social domain of the ADOS contributed to detecting a low FIQ, while the repetitive behavior domain of the ADI-R contributed to detecting a high FIQ. In teenager females, the communication and repetitive behavior domains of the ADI-R as well as the communication domain of the ADOS were highly associated with detecting high FIQ. Interestingly, the domains of the ADI-R and ADOS were not informative of the patients’ level of FIQ in adult males.

Across analyses, our results show that the communication domain of the ADOS was the most salient overall domain in distinguishing sex, age and FIQ (7 contributions) and the ADOS domains were the most informative about the patient’s sex.

## DISCUSSION

Our study emphasizes the relatively bigger importance of certain parts of two widely administered clinical instruments for autism. The social and communication domains of the ADI-R were found particularly informative for predicting autism severity when looking for additive effects as well as non-linear interactions confirming the informative value of these domains in our sample. Further, 3 dimensional partitions of discrete autism profiles were uncovered and these 3 subgroups exhibited the highest discrepancy on the social and communication domains of the two instruments. Additionally, we provide a comprehensive characterization of how information each instrument domain carries about the patients’ age, sex, and FIQ. Collectively, our results provide quantitative insights on the relation between ADI-R and ADOS by using a set of modern data-analysis tools.

### Extracting the most predictive domain subsets in the ADI-R and ADOS

As a primary focus of the present investigation, we algorithmically identified the most predictive ADI-R and ADOS domains of autism severity. Using all domains from the ADI-R and ADOS, the sparse logistic regression predicted autism severity with an accuracy of 97%. In particular, the first solution that gave non-zero coefficient was a subselection of 2 domains which predicted autism severity with an accuracy of about 89% thus only 8% below the accuracy obtained using all domains. The specific subset included the social and communication domains of the ADI-R. This core subset of instrument domains was highly predictive of autism severity.

More generally, the ADI-R and ADOS assessments are often mentioned to be well designed and comprehensive, but not to be particularly efficient to administer in clinical practice. The cost-effectiveness has rarely been carefully evaluated. In fact, ∼45 minutes are required for the ADOS assessment, while approximately ∼1.5 hours are required for the ADI-R assessment. Furthermore, extensive training is needed to use each instrument, which makes the combined assessment very heavy and time-expensive. Berument and colleagues (1999) found that the diagnostic accuracy of the ADI and a 40-item scale was pretty much the same suggesting that much shorter and highly effective screening instrument can be established. In a similar spirit, our results show that assessing only 2 domains from the ADI-R assessment may be sufficient for making reliable statements about the rough overall severity of the psychopathology of patients with autism. In sum, although the combination of ADI-R and ADOS assessments has been shown to improve autism diagnostic validity, we found support for a subset highly predictive of autism severity in a data-guided fashion nevertheless. In other words, using a shorter version of the ADI-R in addition to clinical expertise might be an appropriate tradeoff to alleviate the time constraint.

Mazefski and colleagues (2013) highlighted a discrepancy in the ability of the ADOS to capture autism symptoms catalogued in the DSM-5. These authors found that the ADI-R was more relevant than the ADOS for tapping on the breadth of autism symptoms as defined by the DSM-5. Similarly, Wiggins and Robins (2008) reported that using only the social and communication domains of the ADI-R resulted in improved sensitivity and specificity of the instrument. Our results lend support to these previous findings in showing that the social and communication domains of the ADI-R were more predictive of autism severity than any other domains from the ADOS and therefore potentially more relevant for a fast and effective screening of autism severity.

In sum, our results lend support to the predictive value of the derived instrument subset, comprising the social and communication domains of the ADI-R. From a clinical perspective, being able to find evidence for repetitive behavior has often been considered as being indicative of an autism diagnosis, while only finding interaction and communication difficulties is considered less convincing (Szatmari et al., 2006). This stands in contrast to our empirical findings that emphasizes the predicting value of the interaction and communication domains. This observation adds further arguments in favor of assisting medical decision making in psychiatry by predictive machine learning algorithm towards a future of precision medicine.

### Extracting patient subgroups from the ADI-R and ADOS

Given that autism is widely acknowledged to be a spectrum disorder, we aimed at providing windows into useful intermediate phenotypes. Using a clustering algorithm, three distinct types of clinically meaningful symptom sets emerged: a severe profile with high expression in each domain of the ADI-R and ADOS, ii) a mild profile with low scores on both instruments with particularly low associations with the social and communication domains of the ADI-R and iii) an ADOS-negative profile with high scores only in the domains of the ADI-R. In each subgroup, the social and communication domains of both instruments were the most informative markers while the repetitive behavior domain of both instruments was not as informative. Our results provide data-driven evidence that a major difference between autism patients is the extent of the social and communication domain symptoms as provided by the ADI-R and the ADOS. Our analysis also revealed a group of patients scoring high only in the ADI-R. This subgroup indicates that for certain patients, symptoms might remain hidden when only the ADOS is used for the symptom severity assessment. These patients are sometimes classified as ADOS-negative (Philip et al., 2010). This ADOS-negative subgroup suggests that the ADI-R might be a more appropriate tool to accurately capture autism severity when assessing symptoms using both instruments is not achievable. Finally, our results provide hints for the possible clinical effectiveness of the subtypes.

A few existing studies also applied a clustering method to extract meaningful information but only from either the ADOS or the ADI-R. For instance, using K-means clustering, Cholemkery and colleagues (2016) found 3 clusters in a sample of patients assessed with the ADI-R. Their results indicated a different pattern of social interaction and communication problems versus stereotype behaviors across the identified subgroups. The characteristics of the extracted clusters fit with the assumption of a severity gradient across the 3 subgroups they found. Similarly, a severity gradient was the main difference between the subgroups that emerged in our sample. Spiker and colleagues (2002) also used K-means clustering in a sample of siblings with autism and found subgroups that could be characterized along a single and continuous severity dimension. Children with the lowest nonverbal IQ scores exhibited higher scores in each domain of the ADI-R. Those with higher nonverbal IQs had ADI-R scores indicative of less severe impairment. Interestingly, these authors found that repetitive behavior scores were negatively correlated with this severity gradient in their sample which corroborate our results in two ways. First, we also found a severity gradient across the three clusters mostly related to the social and communication domains of the two instruments. Given the ADOS-negative subgroup, the ADI-R social and communication domains might therefore be a good tradeoff to capture symptoms severity if the time constraint discourages the use of the two instruments. Second, in the three subgroups that emerged from our sample, the repetitive behavior domain of the ADI-R and ADOS expressed in the three scenarios an opposite pattern of the severity gradient. In the subgroup exhibiting the highest scores in each domain of the ADI-R and ADOS, the repetitive behavior domain had the lowest weight compared to the remaining domains of the same instrument. In the subgroup exhibiting the lowest scores in each domain, the repetitive behavior domain of the ADI-R and ADOS had the highest one compared to the two others of the same scale.

In sum, our results corroborate previous findings on potential autism subtypes. Furthermore, our results have repeatedly emphasized relevance of the social and communication domains of the ADI-R, which were found to be highly informative of autism severity in our previous analyses.

### Relation of instrument domains to patients’ age, sex, and FIQ

We explored the contribution of each instrument domain in deriving age, sex and level of FIQ of patients in our sample. Our results suggest that i) the ADI-R and ADOS are differently useful to uncover relevant information of patients with autism, and that ii) sex differences might be more contrasting through the ADOS rather than through the ADI-R domains.

Some studies identified differences between girls and boys in social symptoms on the ADI-R (Beggiato et al., 2017; Carter et al., 2007). However, no sex effects were found when the authors controlled for IQ differences (Banach et al., 2009). Our results complemented these previous findings by suggesting that the ADI-R was not informative about the sex. A few authors have reported greater socio-communication difficulties as captured by the ADOS in females (Carter et al., 2007; Frazier, Georgiades, Bishop, & Hardan, 2014; Hartley & Sikora, 2009). Corroborating these results, we found the social and communication domains of the ADOS to be often informative about the patient’s sex. Scoring high in the social domain of the ADOS was indicative of a female in teenagers and adults which contrasts with the “adolescent emergence hypothesis”. This theory states that the social deficits in females appear later in time (Bargiela, Steward, & Mandy, 2016; Mandy, Pellicano, St Pourcain, Skuse, & Heron, 2018). Sex differences should hence appear to be lower as patients grow up which was not the case in our sample. Given that in subsets of adult patients, social and communication domains were as informative about sex differences as in subsets of teenagers, our results are also in contrast with the “female compensation hypothesis”. According to this theory, females learn over time to compensate for their social difficulties (i.e. social camouflage) more effectively than males (Dworzynski, Ronald, Bolton, & Happé, 2012; Lai et al., 2017; Lehnhardt et al., 2016; Mandy et al., 2018) which would lead to a higher sex difference later in time. Interestingly, scoring high in the communication and social domains of the ADOS was indicating of a female in subsets of patients that were teenagers. Thus, our results suggest that social and communication symptom deficits are less visible in teenager male patients which, again, is in contrast with the two previous hypotheses.

Our results shed light on many sex differences in the ADOS scoring. Surprisingly, the ADI-R was little relevant to differentiate patients’ sex in our sample. However, the two instruments were informative about patients’ level of FIQ and age. Indeed, interesting patterns emerged regarding the level of FIQ and age of patients. For example, we found the repetitive behavior domain of the ADI-R to be very useful to distinguish the age and the FIQ level of the patients. A patient scoring high on the repetitive behavior domain of the ADI-R was more likely to be identified as a teenager rather than an adult in every subset of patients. This pattern potentially indicates that these symptoms attenuate with age which was also found in previous studies (Esbensen, Seltzer, Lam, & Bodfish, 2009; Leekam, Prior, & Uljarevic, 2011; Seltzer et al., 2003). Conversely, we found that patients who scored high in the communication domain of the ADOS were more likely to be adults rather than teenagers. These results suggest that communication problems are harder to disguise as patients grow up. In contrast, previous findings from Seltzer and colleagues (2003) found no difference between teenager and adult in the communication symptoms severity.

In sum, our last set of analyses corroborates previous findings showing that the ADI-R and the ADOS are both differently informative about the patients’ age and level of FIQ. Our results also emphasize the higher relevance of the ADOS than the ADI-R to estimate patients’ sex. Finally, our results show that disease-specific patterns of ADI-R and ADOS scores related to patients’ characteristics can be uncovered when studying subset of similar patients’ sex, age or level of FIQ.

### Conclusion

Improving the process of detecting and monitoring autism is an emerging agenda to improve early intervention and reduced cost. On the one hand, our quantitative investigations expose a subset of the ADI-R domains to be sufficient for making reliable statements about the overall severity of autism and uncovered three types of distinct, clinically meaningful patient categories. On the other hand, the discovered subgroup effects showed that patterns of ADI-R and ADOS scores discriminate patients’ sex, age and level of FIQ. A combination of ADI-R and ADOS assessments improve diagnostic validity but worsen the already heavy process to detect autism severity. In today’s medical practice, clinical centers often use only one instrument but the choice of preferred instrument remains quite variable. Our data-driven research identified some principles that can help guide the choice of a most suitable instrument when the time constraint prompts the medical team to use only one. Furthermore, our study paves the way to reconciling and integrating the ADI-R and ADOS. Our repository is a large sample but this participant sample may be limited to its demographic characteristics. Future studies could provide further insights into underlying patterns of the ADI-R and ADOS specific to individuals with autism with lower intellectual ability or younger. It is also important to note that our results on female patients are only preliminary and need to be confirmed by future research. Nevertheless, analysis approaches based on subgroup of patients with similar characteristics may be critical to disentangle the “female compensation” and “adolescent emergence” hypotheses that have been proposed to underlie autism (Bargiela et al., 2016; Beggiato et al., 2017; Dworzynski et al., 2012; Lai et al., 2017; Mandy et al., 2018). Given the heterogeneity among patients, studying subset of patients with similar characteristics can provide advantages on future intervention and research in autism. Increasingly available “big data” resources will allow for such fine-grained explorations.

## Data Availability

Data comes from a publicly available dataset: ABIDE (Autism Brain Imaging Data
Exchange; http://fcon_1000.projects.nitrc.org/indi/abide/). All data analyses were carried out
on this data repository which is described in detail in Di Martino and colleagues (2014)

## BIBLIOGRAPHY

Banach, R., Thompson, A., Szatmari, P., Goldberg, J., Tuff, L., Zwaigenbaum, L., & Mahoney, W. (2009). Brief report: Relationship between non-verbal IQ and gender in autism. Journal of autism and developmental disorders, 39(1), 188.

Bargiela, S., Steward, R., & Mandy, W. (2016). The experiences of late-diagnosed women with autism spectrum conditions: An investigation of the female autism phenotype. Journal of autism and developmental disorders, 46(10), 3281–3294.

Beggiato, A., Peyre, H., Maruani, A., Scheid, I., Rastam, M., Amsellem, F., … Gillberg, C. (2017). Gender differences in autism spectrum disorders: divergence among specific core symptoms. Autism Research, 10(4), 680–689.

Berument, S. K., Rutter, M., Lord, C., Pickles, A., & Bailey, A. (1999). Autism screening questionnaire: diagnostic validity. The British Journal of Psychiatry, 175(5), 444–451.

Bzdok, D. (2017). Classical statistics and statistical learning in imaging neuroscience. Frontiers in neuroscience, 11, 543.

Bzdok, D., & Meyer-Lindenberg, A. (2017). Machine learning for precision psychiatry: Opportunites and challenges. Biological Psychiatry: Cognitive Neuroscience and Neuroimaging.

Carter, A. S., Black, D. O., Tewani, S., Connolly, C. E., Kadlec, M. B., & Tager-Flusberg, H. (2007). Sex differences in toddlers with autism spectrum disorders. Journal of autism and developmental disorders, 37(1), 86–97.

Charrad, M., Ghazzali, N., Boiteau, V., Niknafs, A., & Charrad, M. M. (2014). Package ‘NbClust’. Journal of Statistical Software, 61, 1–36.

Cholemkery, H., Medda, J., Lempp, T., & Freitag, C. M. (2016). Classifying autism spectrum disorders by ADI-R: subtypes or severity gradient? Journal of autism and developmental disorders, 46(7), 2327–2339.

De Bildt, A., Sytema, S., Ketelaars, C., Kraijer, D., Mulder, E., Volkmar, F., & Minderaa, R. (2004). Interrelationship between autism diagnostic observation schedule-generic (ADOS-G), autism diagnostic interview-revised (ADI-R), and the diagnostic and statistical manual of mental disorders (DSM-IV-TR) classification in children and adolescents with mental retardation. Journal of autism and developmental disorders, 34(2), 129–137.

Di Martino, A., Yan, C.-G., Li, Q., Denio, E., Castellanos, F. X., Alaerts, K., … Dapretto, M. (2014). The autism brain imaging data exchange: towards a large-scale evaluation of the intrinsic brain architecture in autism. Molecular psychiatry, 19(6), 659.

Dworzynski, K., Ronald, A., Bolton, P., & Happé, F. (2012). How different are girls and boys above and below the diagnostic threshold for autism spectrum disorders? Journal of the American Academy of Child & Adolescent Psychiatry, 51(8), 788–797.

Esbensen, A. J., Seltzer, M. M., Lam, K. S., & Bodfish, J. W. (2009). Age-related differences in restricted repetitive behaviors in autism spectrum disorders. Journal of autism and developmental disorders, 39(1), 57–66.

Forgy, E. W. (1965). Cluster analysis of multivariate data: efficiency versus interpretability of classifications. biometrics, 21, 768–769.

Frazier, T. W., Georgiades, S., Bishop, S. L., & Hardan, A. Y. (2014). Behavioral and cognitive characteristics of females and males with autism in the Simons Simplex Collection. Journal of the American Academy of Child & Adolescent Psychiatry, 53(3), 329-340. e323.

Frith, U., & Happé, F. (2005). Autism spectrum disorder. Current biology, 15(19), R786–R790.

Goddard, J. A., Lehr, R., & Lapadat, J. C. (2000). Parents of children with disabilities: Telling a different story. Canadian Journal of Counselling and Psychotherapy/Revue canadienne de counseling et de psychothérapie, 34(4).

Goin, R. P., & Myers, B. J. (2004). Characteristics of infantile autism: Moving toward earlier detection. Focus on Autism and Other Developmental Disabilities, 19(1), 5–12.

Goin-Kochel, R. P., Mackintosh, V. H., & Myers, B. J. (2006). How many doctors does it take to make an autism spectrum diagnosis? Autism, 10(5), 439–451.

Grantham, C., Gower, M., McCalla, M., Harris, A., O’Kelley, S., & Guest, K. (2014). Diagnosis of autism utilizing the ADOS and ADI-R: are there factors to account for discrepancies. Paper presented at the International Meeting for Autism Research. San Diego, California Available: https://imfar.confex.com/imfar/2011/webprogram/Paper7956.html. Accessed.

Hartley, S. L., & Sikora, D. M. (2009). Sex differences in autism spectrum disorder: an examination of developmental functioning, autistic symptoms, and coexisting behavior problems in toddlers. Journal of autism and developmental disorders, 39(12), 1715.

Hastie, T., Tibshirani, R., & Wainwright, M. (2015). Statistical learning with sparsity: the lasso and generalizations: CRC press.

Kim, S. H., & Lord, C. (2012). Combining information from multiple sources for the diagnosis of autism spectrum disorders for toddlers and young preschoolers from 12 to 47 months of age. Journal of Child Psychology and Psychiatry, 53(2), 143–151.

Lai, M.-C., Lombardo, M. V., Ruigrok, A. N., Chakrabarti, B., Auyeung, B., Szatmari, P., … Consortium, M. A. (2017). Quantifying and exploring camouflaging in men and women with autism. Autism, 21(6), 690–702.

Leekam, S. R., Prior, M. R., & Uljarevic, M. (2011). Restricted and repetitive behaviors in autism spectrum disorders: a review of research in the last decade. Psychological Bulletin, 137(4), 562.

Lehnhardt, F.-G., Falter, C. M., Gawronski, A., Pfeiffer, K., Tepest, R., Franklin, J., & Vogeley, K. (2016). Sex-related cognitive profile in autism spectrum disorders diagnosed late in life: implications for the female autistic phenotype. Journal of autism and developmental disorders, 46(1), 139–154.

Lemler, M. (2012). Discrepancy between parent report and clinician observation of symptoms in children with autism spectrum disorders. Discussions, 8(2).

Lord, C., Risi, S., Lambrecht, L., Cook, E. H., Leventhal, B. L., DiLavore, P. C., … Rutter, M. (2000). The Autism Diagnostic Observation Schedule—Generic: A standard measure of social and communication deficits associated with the spectrum of autism. Journal of autism and developmental disorders, 30(3), 205–223.

Lord, C., Rutter, M., & Le Couteur, A. (1994). Autism Diagnostic Interview-Revised: a revised version of a diagnostic interview for caregivers of individuals with possible pervasive developmental disorders. Journal of autism and developmental disorders, 24(5), 659–685.

Mandy, W., Pellicano, L., St Pourcain, B., Skuse, D., & Heron, J. (2018). The development of autistic social traits across childhood and adolescence in males and females. Journal of Child Psychology and Psychiatry.

Mazefsky, C., McPartland, J., Gastgeb, H., & Minshew, N. (2013). Brief report: Comparability of DSM-IV and DSM-5 ASD research samples. Journal of autism and developmental disorders, 43(5), 1236–1242.

Mazefsky, C. A., & Oswald, D. P. (2006). The discriminative ability and diagnostic utility of the ADOS-G, ADI-R, and GARS for children in a clinical setting. Autism, 10(6), 533–549.

Philip, R., Whalley, H., Stanfield, A., Sprengelmeyer, R., Santos, I., Young, A., … Lawrie, S. (2010). Deficits in facial, body movement and vocal emotional processing in autism spectrum disorders. Psychological medicine, 40(11), 1919–1929.

Quine, L., & Pahl, J. (1987). First diagnosis of severe handicap: a study of parental reactions. Developmental Medicine & Child Neurology, 29(2), 232–242.

Seltzer, M. M., Krauss, M. W., Shattuck, P. T., Orsmond, G., Swe, A., & Lord, C. (2003). The symptoms of autism spectrum disorders in adolescence and adulthood. Journal of autism and developmental disorders, 33(6), 565–581.

Shalev-Shwartz, S., & Ben-David, S. (2014). Understanding machine learning: From theory to algorithms: Cambridge university press.

Spiker, D., Lotspeich, L. J., Dimiceli, S., Myers, R. M., & Risch, N. (2002). Behavioral phenotypic variation in autism multiplex families: evidence for a continuous severity gradient. American Journal of Medical Genetics, 114(2), 129–136.

Szatmari, P., Georgiades, S., Bryson, S., Zwaigenbaum, L., Roberts, W., Mahoney, W., … Tuff, L. (2006). Investigating the structure of the restricted, repetitive behaviours and interests domain of autism. Journal of Child Psychology and Psychiatry, 47(6), 582–590.

Wiggins, L. D., & Robins, D. L. (2008). Brief report: Excluding the ADI-R behavioral domain improves diagnostic agreement in toddlers. Journal of autism and developmental disorders, 38(5), 972–976.

